# Device sensitivity and false alarms can reshape regression-to-the-mean in simulated epilepsy trials

**DOI:** 10.64898/2026.07.30.26359361

**Authors:** Daniel M. Goldenholz, Shira R. Goldenholz, Rohan Bhansali, Ted J. Kaptchuk, Brandon Westover

**Affiliations:** Department of Neurology, Beth Israel Deaconess Medical Center, Boston, Massachusetts, USA; Harvard Medical School, Boston, Massachusetts, USA; Program in Placebo Studies, Beth Israel Deaconess Medical Center, Boston, Massachusetts, USA; Department of Neurology, Stanford University, Stanford, California, USA

**Keywords:** seizure detection, clinical trials, regression to the mean, placebo response, seizure diary, false alarm rate

## Abstract

Automated seizure detection devices are increasingly plausible tools for epilepsy trials, but no device is perfect. We used CHOCOLATES, a realistic seizure diary simulator, to examine how device sensitivity and false alarm rate (FAR) affect regression-to-the-mean (RTM) and placebo median percentage change (MPC) in a simulated randomized trial design. For each device condition, 100,000 potential participants were generated; eligibility was assessed during a 2-month baseline, followed by a 3-month test period. With FAR fixed at 0, reducing sensitivity from 100% to 10% increased the fraction of eligible participants exhibiting RTM from 38.2% to 64.8% and increased placebo MPC from 14.7% to 48.1%. With sensitivity fixed at 100% and expected FAR correction, increasing FAR from 0 to 1 alarm/day increased RTM from 38.2% to 53.2% and placebo MPC from 14.7% to 31.3%. Imperfect seizure detection can therefore change the apparent placebo response expected from RTM.

**Short summary for table of contents:** In simulated epilepsy trials, imperfect seizure detection altered regression to the mean and placebo median percentage change. Trial planning should model detector sensitivity and false alarm rate before device-derived seizure counts are used as endpoints.

## Introduction

Automated seizure detection is evolving from an aspirational technology to practical clinical and research use. Current devices are evaluated primarily by sensitivity, false alarm rate (FAR), detection latency, and usability, with recent guidelines and meta-analytic studies emphasizing that performance varies across seizure types and recording settings.^1–3^ These metrics are usually interpreted at the level of patient safety, caregiver notification, or diary accuracy. Their consequences for randomized controlled trial (RCT) design, however, are less developed.

Epilepsy trials are particularly vulnerable to apparent placebo response for several reasons. First, seizure frequency is highly variable over time. Therefore, eligibility criteria often result in patients being included during temporarily high seizure frequency periods. and outcomes are defined as change from baseline.^4–10^ Second, an important component of placebo response is regression-to-the-mean (RTM)^8,11^. Prior work has shown that regression-to-the-mean (RTM) can be manipulated by eligibility criteria and can materially affect placebo response and trial power.^4^ A separate line of work suggests that natural seizure-frequency variability can reproduce many features of placebo response even without a psychobiological placebo effect.^5–8,12^

Seizure detection devices change the measured seizure time-series before trial eligibility and outcomes are calculated. Lower sensitivity can miss true seizures; false alarm rate (FAR) can add false events. Either process could change who qualifies for a trial and the apparent change from baseline. We investigated to what extent sensitivity and FAR alter RTM and placebo response in a simulated epilepsy RCT.

## Methods

We performed a simulation study using CHOCOLATES, an open-source realistic seizure diary simulator that recapitulates observed diary properties, including heterogeneous long-term seizure rates, the relationship between mean seizure frequency and variability, seizure clustering, cycles, and inter-seizure constraints.^12^ Analysis was conducted with custom software in Python 3.13.2.

Each simulated placebo arm participant had a 2-month baseline and 3-month test period. There was no active treatment effect added, consistent with our prior studies showing that placebo response primarily reflects natural variability and RTM^4,12,13^. Eligibility was tested during the baseline period. The baseline criteria were modeled after focal seizure trial eligibility rules used in cenobamate studies: mean baseline frequency at least 4 seizures/month, each baseline month at least 3 seizures, and no seizure-free interval longer than 25 days.^14,15^

Device sensitivity was implemented probabilistically at the daily seizure-count level: each true seizure had probability equal to the specified sensitivity of being detected. FAR was implemented as Poisson-distributed false alarms added to each day. For any analysis involving FAR>0, a correction factor was applied to improve the signal quality from each patient^3^. The expected false alarm count (FAR x duration) was subtracted from seizure counts to improve count estimates. This represents an idealized setting in which device FAR is known and is explicitly accounted for when deriving trial seizure counts.

Two device-performance sweeps were conducted. First, FAR was fixed at 0 alarms/day while sensitivity varied from 10% to 100% in 10% increments. Second, sensitivity was fixed at 100% while FAR varied from 0 to 1 alarm/day in 0.1 alarm/day increments. For each condition, 100,000 potential participants were generated. Among eligible participants, RTM was defined as a baseline seizure frequency above the participant’s effective long-term seizure frequency and closer proximity of the test-period frequency to that long-term frequency. Placebo response was quantified as median percentage change (MPC), with percentage change defined as 100 x (1 - test-period seizure frequency / baseline seizure frequency). Confidence intervals in Figure 1 are nonparametric 95% confidence intervals for the estimated median MPC, calculated from binomial order statistics around the sample median. They are not 95% intervals of individual patient percentage changes. No hypothesis tests were performed, because the analysis was a descriptive simulation sweep rather than a comparison of randomized groups. No missing-data procedure was required because all simulated diaries were complete.

**Figure 1.**
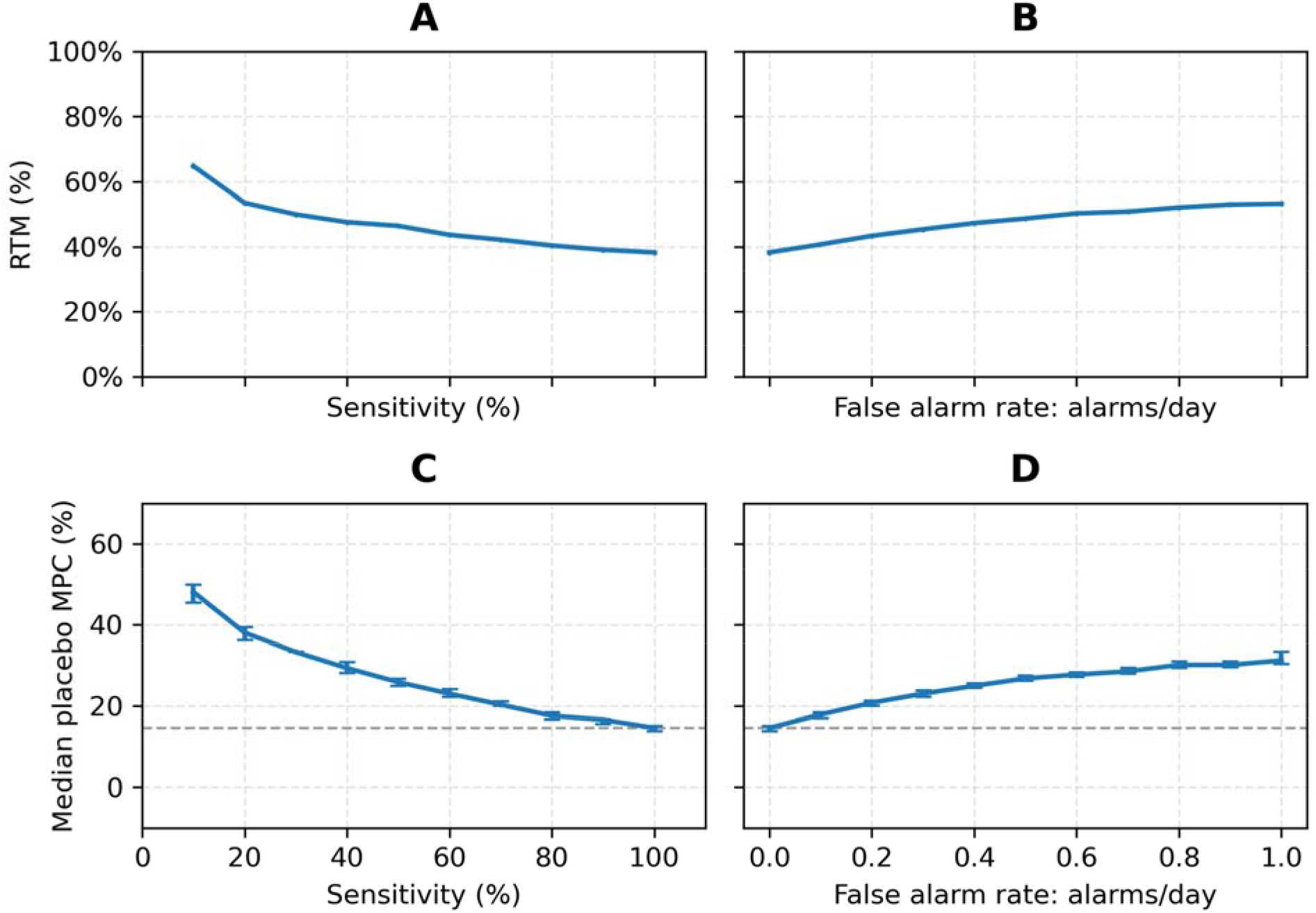
Device sensitivity, false alarm rate (FAR), regression to the mean (RTM), and placebo median percentage change (MPC). Simulations used 100,000 potential participants per condition and baseline-only eligibility. Panels A and C vary sensitivity with FAR fixed at 0 alarms/day. Panels B and D vary FAR with sensitivity fixed at 100%, with expected false alarms subtracted. RTM, regression to the mean; FAR, false alarm rate; MPC, median percentage change. Error bars show 95% confidence intervals for the estimated median MPC.

**Figure 2.**
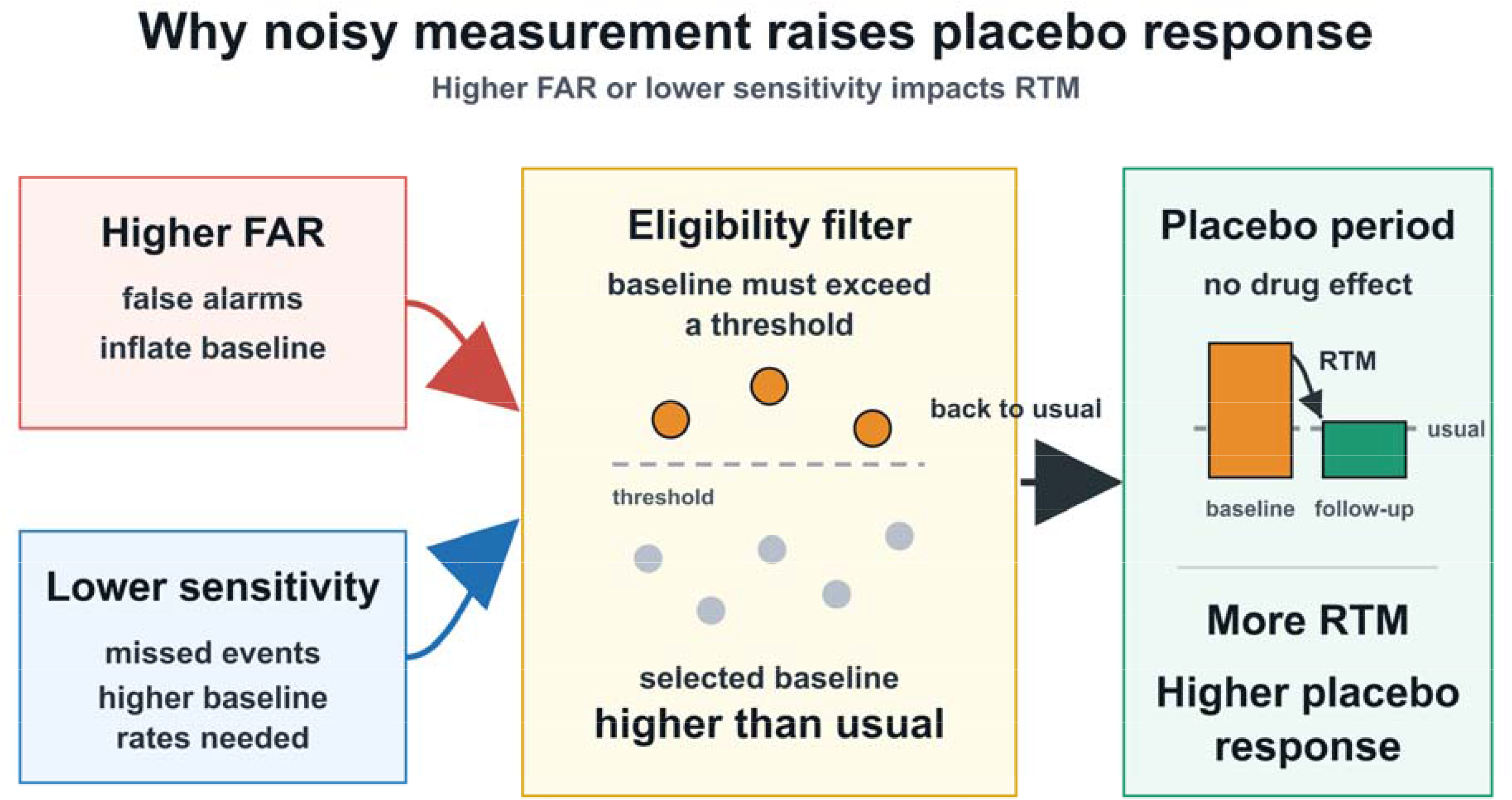
Conceptual mechanism linking detector error to apparent placebo response. Lower sensitivity and higher false alarm rate (FAR) change the measured seizure-count series used for trial eligibility and baseline comparison. Because eligibility selects higher measured baseline seizure counts, detector error can enrich for participants whose later measured counts regress toward their long-term seizure frequency, increasing apparent placebo response.

This was a simulation study using synthetic participants only. No human participants, identifiable private information, or animals were used. The open-source repository is on Github.

## Results

When FAR was 0, lower sensitivity increased both RTM fraction and placebo MPC (Figure 1: A,C). At 100% sensitivity, 33,435 of 100,000 potential participants were eligible, 38.2% of eligible participants met the RTM definition, and median placebo MPC was 14.7% (95% CI 13.9%-15.2%). At 50% sensitivity, 15,724 participants were eligible, RTM was 46.4%, and median placebo MPC was 25.9% (95% CI 25.0%-26.7%). At 10% sensitivity, only 1,660 participants were eligible, but RTM increased to 64.8%, and median placebo MPC increased to 48.1% (95% CI 45.5%-50.0%).

When sensitivity was 100%, increasing FAR also increased RTM and placebo MPC despite correction for expected false alarms (Figure 1: B,D). At FAR 0.1 alarm/day, 35,377 participants were eligible, RTM was 40.7%, and median placebo MPC was 17.9% (95% CI 17.1%-18.5%). At FAR 0.5 alarms/day, 35,522 participants were eligible, RTM was 48.7%, and median placebo MPC was 26.8% (95% CI 26.2%-27.5%). At FAR 1 alarm/day, 35,500 participants were eligible, RTM was 53.2%, and median placebo MPC was 31.3% (95% CI 30.3%-33.3%).

Thus, both missed seizures and false alarms changed the placebo response expected from natural variability and baseline selection alone. The direction was consistent in the primary displayed analysis: lower sensitivity and higher FAR increased the RTM fraction and increased placebo MPC.

## Discussion

This simulation suggests that device performance characteristics should be treated as trial-design parameters, not merely as validation metrics. In a conventional baseline-versus-treatment design, the measured baseline seizure rate determines eligibility and also serves as the denominator for percent-change outcomes^4^. Imperfect detection changes both. Lower sensitivity enriched the eligible cohort for unusually high measured baseline periods among people whose true long-term seizure frequency was lower, producing larger subsequent apparent improvement. FAR had a similar effect even when expected false alarms were subtracted, because false alarms could still alter daily and monthly eligibility behavior and trial selection.

These findings extend prior RTM work in epilepsy RCTs.^4^ The earlier analysis showed that eligibility criteria can increase or decrease RTM and placebo response. The present analysis adds that the measurement system itself can alter the same pathway. If future trials use device-derived seizure counts, then two trials with identical biological treatment effects and identical eligibility rules could have different apparent placebo response solely because the devices or algorithms differ.

A framework for understanding RTM historically suggested two subtypes: RTM type 1, which represented temporary unusual circumstances (such as illness) which upon removal result in return to the usual state, and RTM type 2, which represents the effect of natural variability always going back to the typical state^8^. The results of the present study suggests that the framework should be extended to include RTM type 3, which represents the impact of measurement noise on a system (such as from seizure detectors). Here, we show that RTM type 3 alone can increase placebo response when all other factors are held constant.

Objective seizure detection could reduce under-reporting and may improve statistical power when more complete seizure counts are available.^3,16^ Device operating characteristics should be incorporated into trial simulations before RCT designs are finalized. Prior to initation of the RCT, investigators should specify how sensitivity, FAR, and FAR correction will enter eligibility, baseline counts, outcome counts, and missing-data rules. Device validation should also report performance in the seizure types and clinical settings that match the planned trial population.^1,2^

The limitations of this study must be considered as well. First, CHOCOLATES is a simulator which comes with simplifications of real-world measurements.^12^ Second, sensitivity and FAR were modeled simply and uniformly across participants and time. Real detectors may have patient-specific performance, seizure-type-specific sensitivity, state dependence, clustering of false alarms, periods of device non-wear, or signal deficiency. Third, the FAR correction was idealized. In practice, false alarms may not be known at the individual-event level, and correction rules could affect eligibility and outcomes differently. Fourth, the confidence intervals in Figure 1 quantify uncertainty in the estimated median MPC from simulation, not the dispersion of patient-level responses.

Seizure detection devices may make epilepsy trials more objective. They will also make trial statistics depend on detection algorithm performance in a non-linear fashion. RTM and placebo response should therefore be evaluated jointly with sensitivity and FAR whenever device-derived seizure counts are used for epilepsy RCT eligibility or endpoints.

## Data Availability

The open-source code is available here: https://github.com/GoldenholzLab/RTM3.git.

## Acknowledgments and funding

Daniel M. Goldenholz (DMG) was funded by the National Institutes of Health (NIH) K23NS124656, 1R21NS142800 and the American Board of Psychiatry and Neurology (ABPN). SRG, TJK report no funding. RB reports funding from NIH K23NS124656. OpenAI Codex was used to assist with coding and portions of manuscript preparation; however the authors take full responsibility for the entire project including code, images, and manuscript.

## Disclosure of conflicts of interest

DMG has been provided speaker fees from Harvard Medical School, the American Academy of Neurology, the American Epilepsy Society, the American Clinical Neurophysiology Society, the National Neurotrauma Society, AI in Epilepsy and Neurology, Florida Epilepsy Alliance, and the University of Texas at Austin. He also previously has been a paid consultant for Neuro Event Labs, IDR, LivaNova, Health Advances, Duke University, Bloom Insights, and Wiley. He has received grants from NIH, ABPN, Beth Israel Deaconess Medical Center, and the Lions Club. Rohan Bhansali and Brandon Westover have no relevant conflicts of interest to disclose.

## Ethical publication statement

We confirm that we have read the Journal’s position on issues involved in ethical publication and affirm that this report is consistent with those guidelines.

## Data and code availability

The open-source code is available here: https://github.com/GoldenholzLab/RTM3.git.

## Author contributions

DMG conceived the analysis, wrote and revised analysis code, interpreted results, and drafted the manuscript. RB, SRG, TJK and MBW contributed to interpretation and manuscript revision.

